# Analysis of genome characteristics and transmission of SARS-CoV-2 strains in North-East of Romania during the first COVID-19 outbreak

**DOI:** 10.1101/2020.12.22.20248741

**Authors:** Andrei Lobiuc, Mihai Dimian, Roxana Gheorghita, Olga Sturdza, Mihai Covasa

## Abstract

Romania officially declared its first SARS-CoV-2 case on February 26, 2020. The first and largest COVID-19 outbreak in Romania was recorded in Suceava, N/E region of the country, and originated at the Suceava regional county hospital. Following sheltering-in-place measures, infection rates decreased, only to rise again after relaxation of measures. This study describes the incursion of SARS-CoV-2 in Suceava and other parts of Romania and analyzes the mutations and their association with clinical manifestation of the disease during the period of COVID-19 outbreak. Phylogenetic analysis indicated multiple sites of origin for SARS-CoV-2 strains in Suceava, specifically from Spain, Italy and Russia, but also other strains related to those from Czech Republic, Belgium and France. Most Suceava samples contained mutations common to European lineages, such as A20268G, however aproximately 10% of samples were missing such mutations, indicating a possible different origin. While overall genome regions ORF1ab, S and ORF7 were subject to most mutations, several recurring mutations such as C27707T were identified, and these were mainly present in severe forms of the disease. Non-synonymous mutations, such as C3225A (Thr987Asn in NSP3a domain), associated with changes in a protein responsible for decreasing viral tethering in human host were also present. Patients with diabetes and hypertension exhibited eight and three time,s respectively, higher odds ratios of acquiring severe forms of the disease and these were mainly related to C27707T mutation. These results will aid in tracing virus movement throughout Romania and identification of infectivity, virulence and pathogenicity.

## Introduction

Corona virus disease 2019 (COVID-19) emerged in December 2019 in Wuhan City, Hubei Province, China, in humans exposed to wildlife at the Huanan seafood wholesale market [1]. Provisionally named 2019-nCoV, the International Committee on Virus Taxonomy renamed the virus Severe Acute Respiratory Syndrome Coronavirus-2 (SARS-CoV-2) [2]. Coronaviruses belong to the subfamily Coronavirinae, order Nidovirales and are common human pathogens. They are enveloped, positive-sense RNA viruses with a diameter of 60–140 nm and 29903 base pair single stranded RNA genome. These viruses are characterized by clublike spike projections of protein on the surface, with a crown-like (from the latin”coronam”) appearance under the electronic microscope [3].

The SARS-CoV-2 colonizes the respiratory tract system causing symptoms similar to those of common cold, such as respiratory disorders, runny nose, dry cough, dizziness, sore throat and body aches, headaches and fever for several days [4]. In early stages, patients show acute respiratory infection symptoms, with some quickly developing acute respiratory failure and other serious complications [5]. This virus is transmitted from person to person primarily via aerosolized droplets [6]. To reduce transmission, preventive measures have been recommended, such as mask wearing, frequent hand washing, limiting contact when symptoms are obvious, avoiding public contact and quarantine [7]. Generally, the body’s immune response to SARS-CoV-2 and SARS-CoV is relatively similar and is characterized by an excessive production of cytokines [8].

The first whole genome sequence was published on January 5, 2020, and since then thousands of genomes have been sequenced and deposited in the Global Initiative on Sharing All Influenza Data (GISAID) database [9]. This data revealed that it shares approximately 79.6% similarity with SARS-CoV at the nucleotide level, and varies between the different genes. SARS-CoV-2 contains a linear single-stranded positive-sense RNA as genetic material that encodes for the spike (S), envelop (E), membrane (M), and nucleocapsid (N) proteins [10]. The S glycoprotein is a transmembrane protein found on the viral outer membrane. S protein forms homotrimers that protrude the viral surface and facilitate binding of viral envelope to host cells by interacting with angiotensin-converting enzyme 2 (ACE2) receptors expressed on the lower respiratory tract cells.

Since the introduction of SARS-CoV-2 in territories outside Asia, continous efforts have been made to map strains and lineages. With time, the viral spread brought about mutations specific to geographical regions, thus making possible to track the virus movement within communities and across the globe. One of the most well known mutations is in position 23404, changing an aspartate for a glycine at residue 614 in Spike protein and, presumably, offering an advantage in viral replication [11]. This mutation appeared in January 2020 in China, and after a week in Europe and, was later observed in Africa and Americas [12], giving birth to the “G” clade, now characteristic to Europe [13]. In other geographical settings, USA samples share mutations at positions 8782, 17747, 17858, 18060, and 28144, with the first and the latter also present in European samples. Such signatures, composed of multiple reccurent mutations within the same region have been considered, when identifying founder effects for that lineage [14].

Identifying mutations and strains movement across geographical regions is critical for predicting further infection hotspots, as well as for vaccine and diagnostic tests development. The first patient with COVID-19 in Romania was confirmed on February 26, 2020, in Gorj county, South-West of Romania [15]. On March 3, 2020, pacient number 6 was diagnosed with COVID-19 and hospitalized at the Regional County Hospital of Suceava, North East of Romania. This led to a rapid contamination of medical personnel and the Regional Hospital of Suceava became the largest outbreak of COVID-19 in the country that still leads in the number of confirmed cases and deaths nationwide. Suceava Regional Hospital serves more than 600,000 people and is the largest hospital in the North-East region of the country. On March 26, a SARS-CoV-2 diagnostic laboratory (RT-PCR based) was set up within the hospital that allowed identification of pacients with SARS-CoV-2 infection. Suceava county has one of the largest migrant population working in EU countries who begun returning to Romania once COVID-19 spread accross Europe. The initial epidemiological analysis on a small sample of patients from Bucharest (capital of Romania) and several counties collected between February and March 2020 indicated that Romanian migrants from Italy were the main source of virus spread [16]. In order to understand the source and transmission of the virus of this largest outbreak, we perfomed sequencing and phylogenetic analyses of viral samples from patients with confirmed SARS-CoV-2 infection from Suceava county. We then compared the data with those reported from other regions of the country as well as several European countries accessed from GISAID data. Finally, we examined whether specific variants in SARS-CoV-2 proteins were associated with patients’ clinical parameters and disease outcomes.

## Results

### Characteristics of sequenced samples from Suceava

A total of 62 samples were selected, based on quality checks, comprising 39 samples from males and 23 from females. When compared with Wuhan reference Genome, GISAID accesion ID Nc_045512.2, a total of 190 modifications were recorded and distributed across 8 genome regions. With ORF1ab being the largest SARS-CoV-2 gene (approximately 24 kb), corresponding to a polyprotein made up of 16 non-structural proteins (NSP1-16), over 66% of all mutations were recorded in this region. This was followed by the spike (S) and nucleocapsid (N) protein coding genes while other genes, such as ORF3a, ORF7a or envelope (E) represented less than 5% of all mutations (Fig. 1).

**Fig 1.**
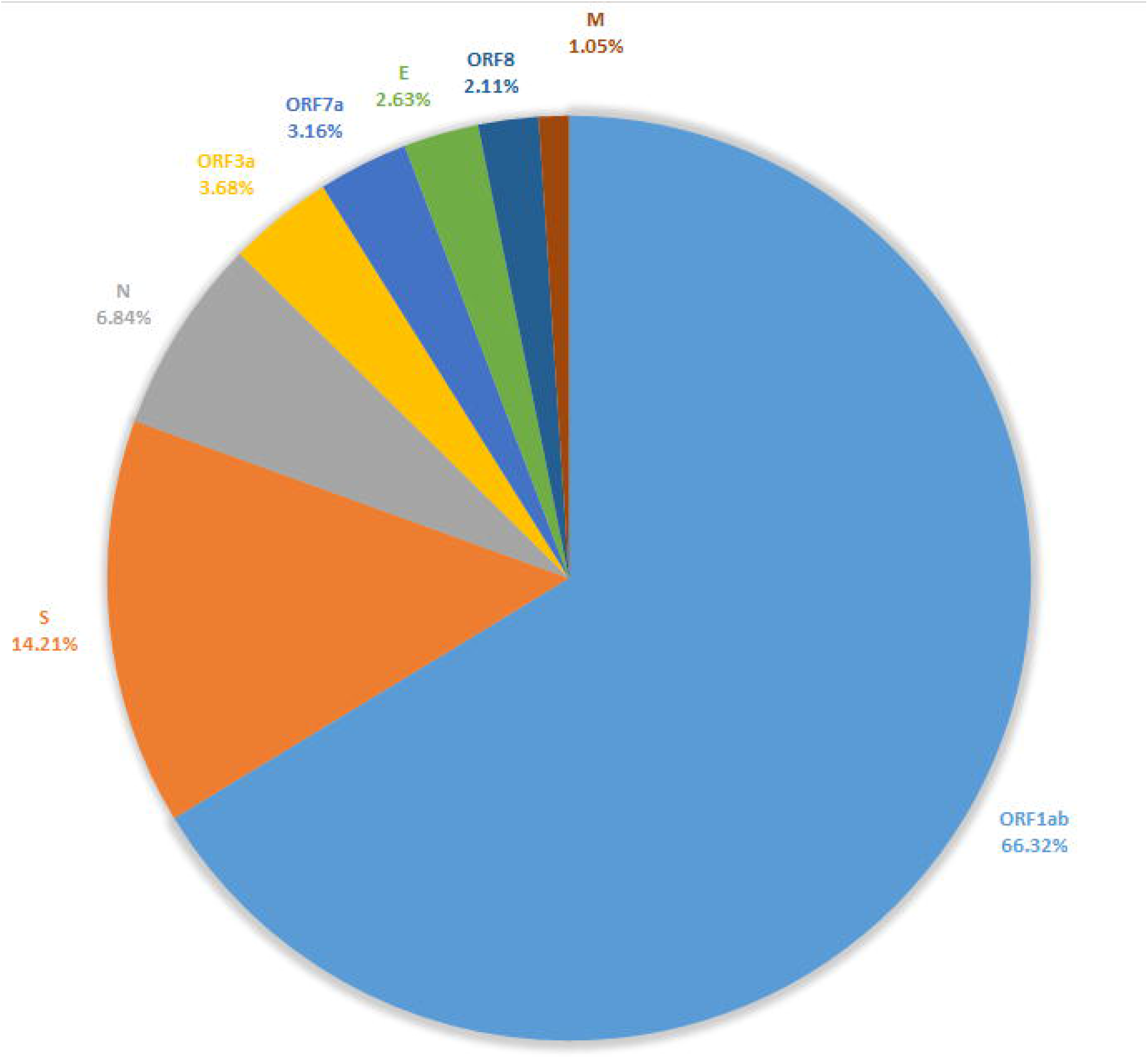
Percentages of nucleotide modifications, distributed over main genome regions, compared to Wuhan-Hu1 reference strain. When observing instances of individual mutations, rather than gene-wise frequencies, most frequent modified nucleotides were recorded at positions 241, 3037, 14408, 23403, 20268, 27707, and 9697, totaling 57% of all modifications. These mutations were present in as few as 5, and up to all 62 samples. A distribution of recurrent modifications throughout the genome (∼30 kb) is shown in Fig. 2.

**Fig 2.**
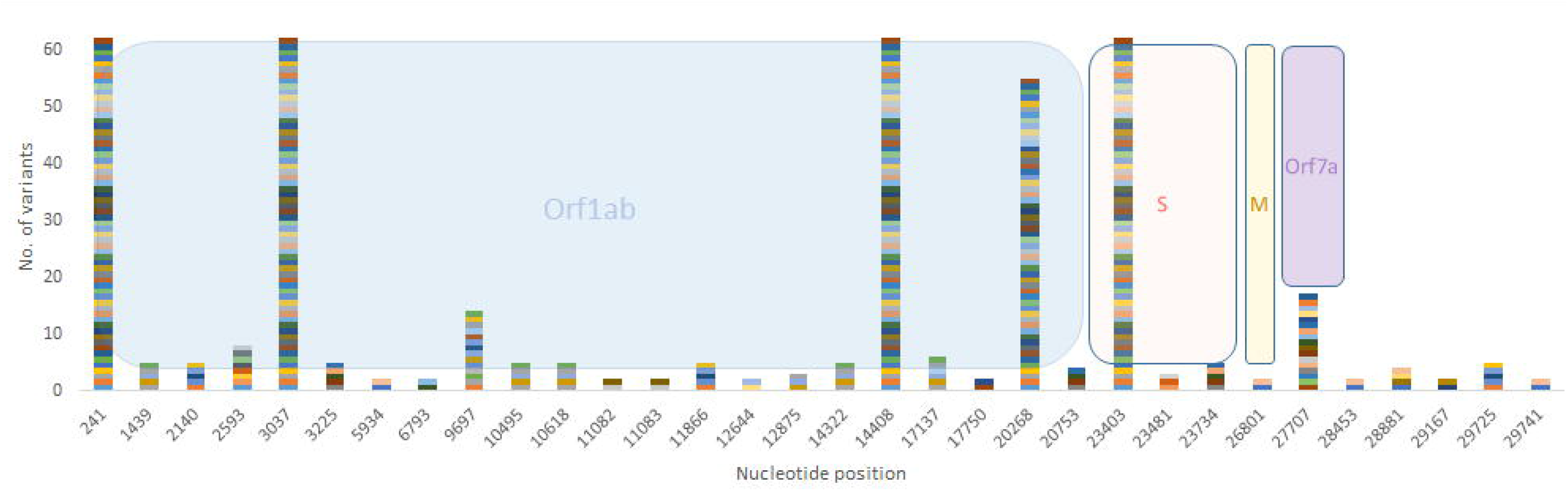
Most frequently modified nucleotide positions and corresponding genome regions. Of the mutations recorded, the highest proportion belongs to transitions, accounting for over 76% of them, while only 23.7 % are transversions. Specifically, C to T (U) transitions make up almost 50% of all SNPs recorded, followed by G to T (U) transversions (17%), A to G (11%) and T(U) to C (10%) transitions (Fig. 3). Overall, the transitions:transversions ratio is 3.2:1, with a value >1 for most genes.

**Fig 3.**
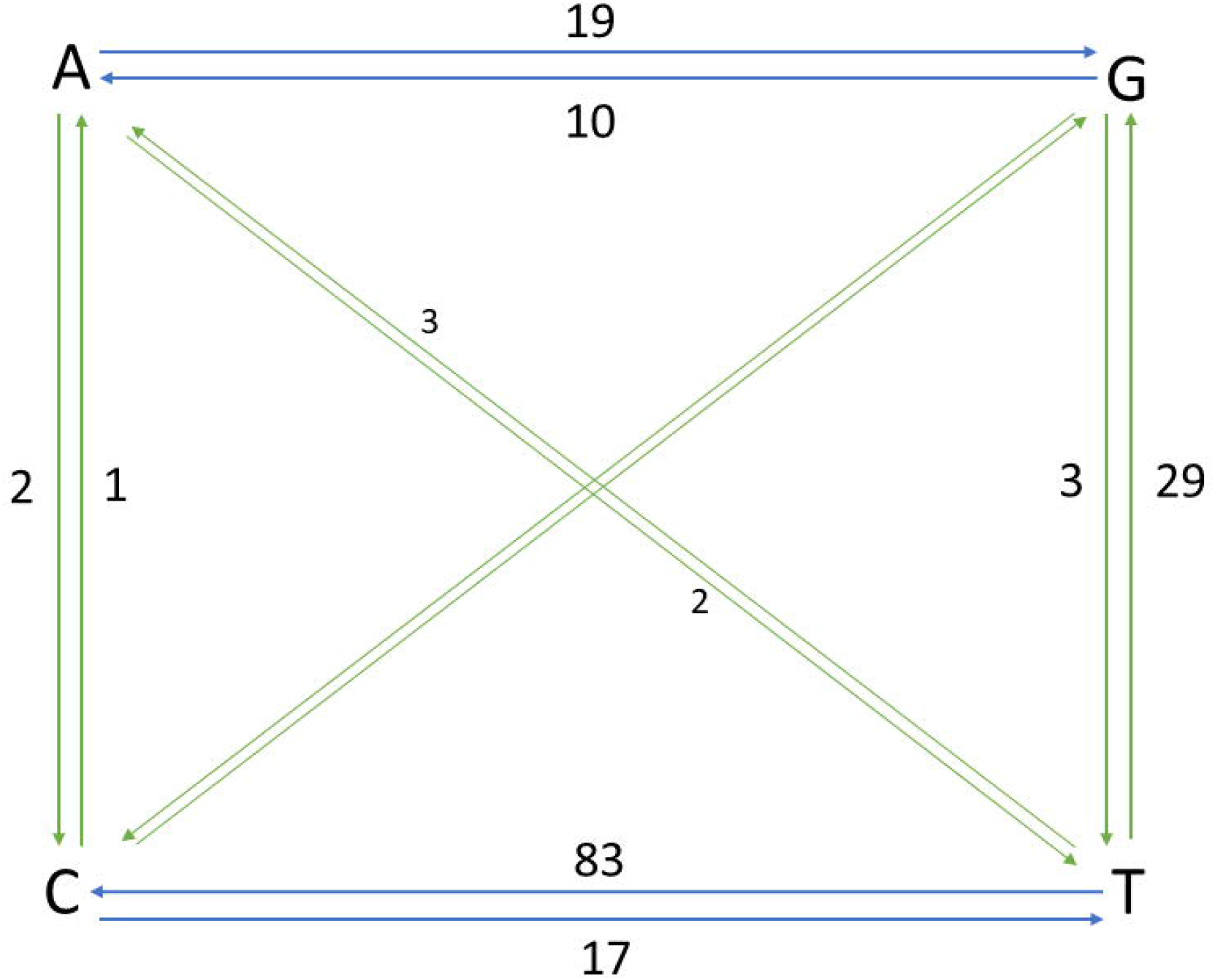
Base changes in analysed sequences. Each stacked color indicates a mutation at the corresponding position within the genome region designated in the colored areas. Numbers next to arrows indicate how many times that particular mutation was recorded; T is replaced by U in the original, RNA sequence; blue, transitions; green, transversions.

When considering aminoacid sequences, modifications at most frequent positions were of synonymous type, however, a large proportion induced aminoacid alterations, such as those at positions 314 in ORF7a and 1841 in S genes (Table 1). At the entire genome level, for certain ORFs, the number of non-synonymous mutations was higher than that of synonymous ones (Fig. 4).

**Table 1.** Frequent nucleotide and aminoacid modifications

| Nucleotide position | Genome region | Position within gene/Nucleotide modification | Aminoacid modification | Type of modification |
| --- | --- | --- | --- | --- |
| 3037 | ORF1ab | 2772C>T | p.Phe924Phe | synonymous_variant |
| 14408 | ORF1ab | 14143C>T | p.Leu4715Leu | synonymous_variant |
| 23403 | S | 1841A>G | p.Asp614Gly | missense_variant |
| 20268 | ORF1ab | 20003A>G | p.Ter6668Trpext*? | stop_lost |
| 27707 | ORF7a | 314C>T | p.Ala105Val | missense_variant |
| 9697 | ORF1ab | 9432T>C | p.Tyr3144Tyr | synonymous_variant |

**Fig 4.**
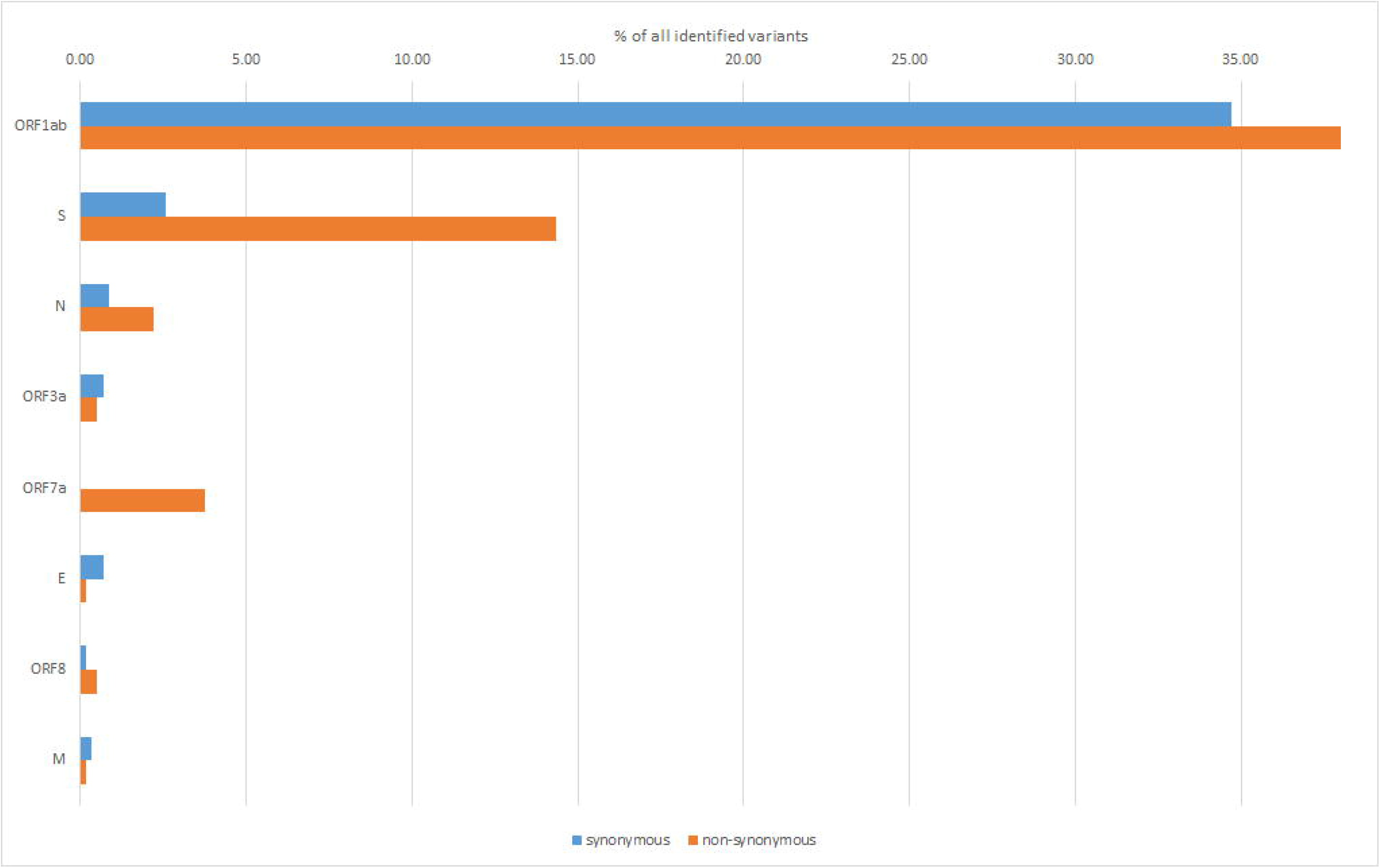
Percentages of genome-wide synonymous/non-synonymous variants. The well known S-protein mutation, D614G that gives rise to GISAID “G” clade, was present in all 62 samples. Other key signatures of SARS-CoV-2 evolution were changes at positions 14408 and 20268. A notable feature was the mutation at position 27707, leading to a change from alanine to valine in ORF7a sequence, at position 105 and present in 17 out of the 62 analysed samples.

The majority of nucleotide modifications preserved the reading frames of the genetic sequences, however, a number of deletions led to gene variants, including frameshift ones (Table 2). Out of these, S gene modification, in position 29725, was present in 5 out of 62 analysed samples (8 %).

**Table 2.**
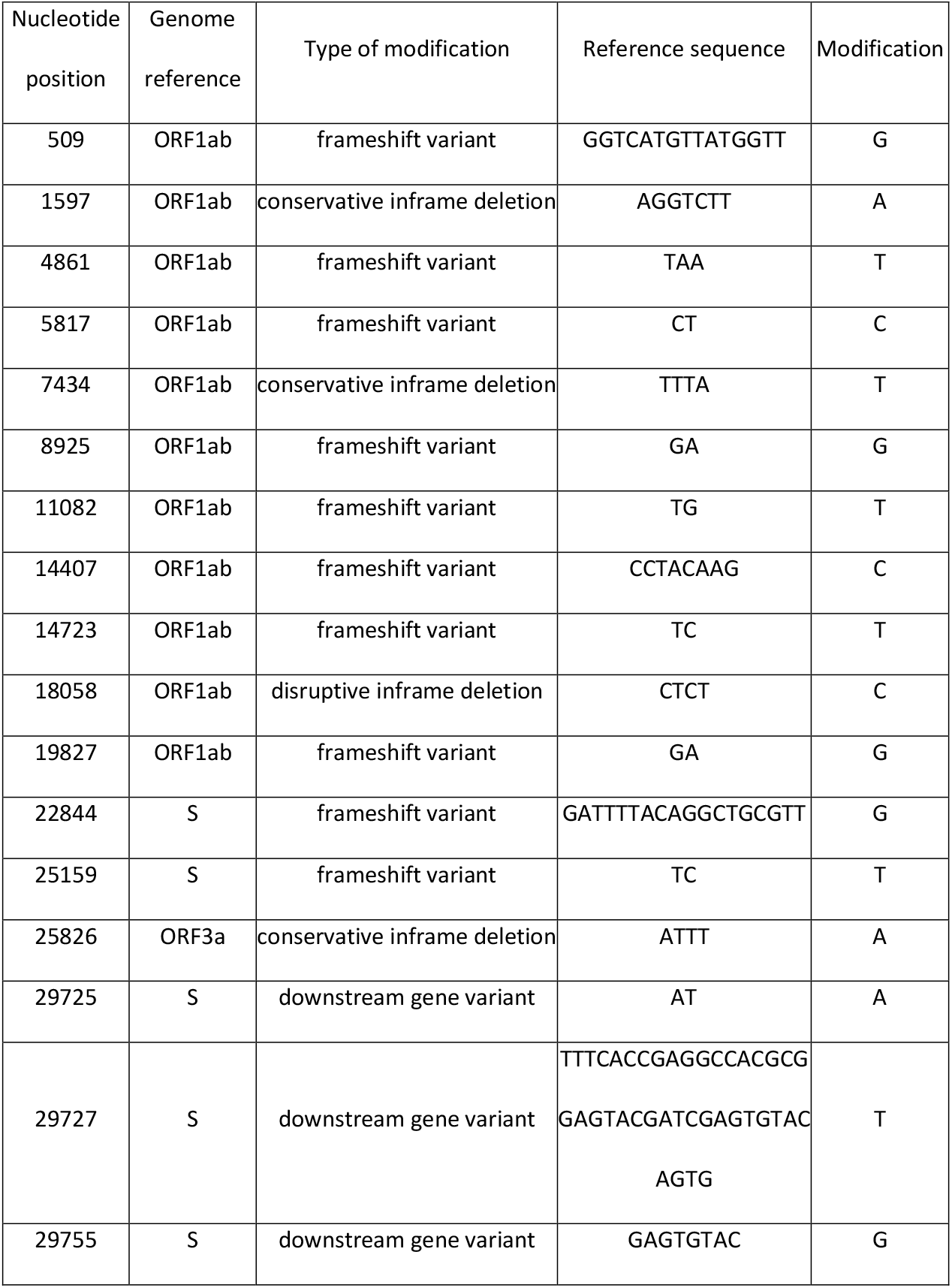
Nucleotide modifications leading to gene variants

### Phylogenetic analyses

Suceava outbreak was the first and the largest in the country. The evolution of SARS-CoV-2 infection was chronologically different in Suceava, compared to other regions of the country, peaking in March then decreasing sharply while in the rest of the country, particularly in the N-E region there was a gradual increase in case prevalence (Fig. 5).

**Fig 5.**
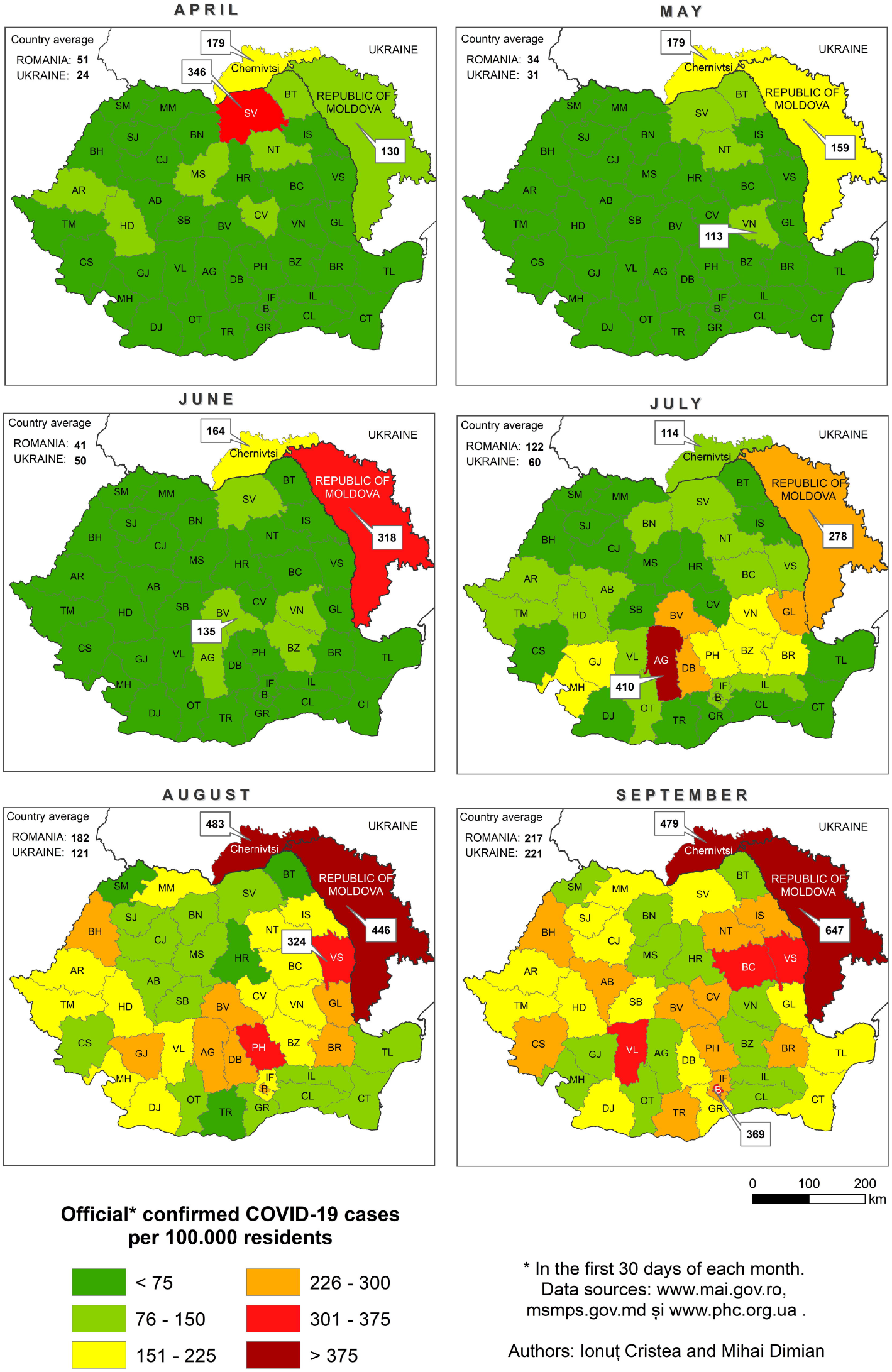
The evolution of SARS-CoV-2 infections in Suceava (SV) and the rest of Romania, during April-September 2020. To examine the phylogenetic distribution of SARS-CoV-2 from Suceava county, we first compared these samples (n=62) with those from other regions of Romania (n=50). All sequences were submitted to GISAID (https://www.gisaid.org/) with accession numbers listed in S1 Table. The total number of samples (n=112) were grouped in 6 clusters, 4 of which included samples from Suceava. Most Suceava samples (n=45) were grouped in a large cluster, together with few strains from Bucharest. A smaller number of sample sequenced in our laboratory (n=12) belonged to another distinct cluster with samples from Bucharest. Four samples clustered with those from Bucharest and Buzau and one sample was closer to a strain from Iasi (Fig. 6).

**Figure 6.**
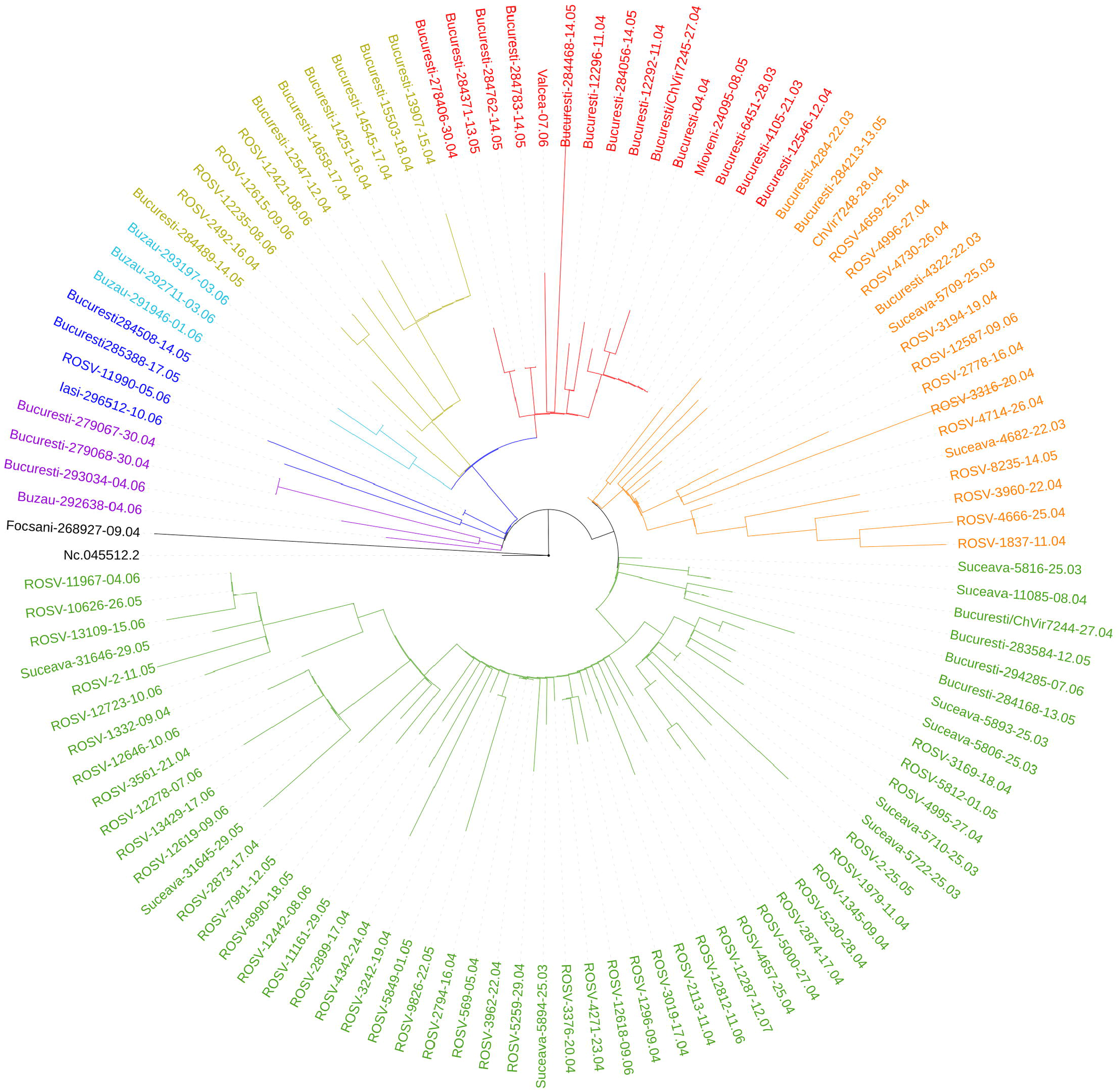
Phylogenetic distribution of Romanian strains (n=112). Then, we compared phylogenetic distribution of Romanian strains with representative samples from European viral genomes. To do this, a maximum likelihood tree was constructed, using bootstrapped RaxML (Fig. 7). Overall, Romanian strains formed several clusters with strains from specific European regions such as Spain, Russia, Italy, Turkey, England and Austria (Fig. 7).

**Fig 7.**
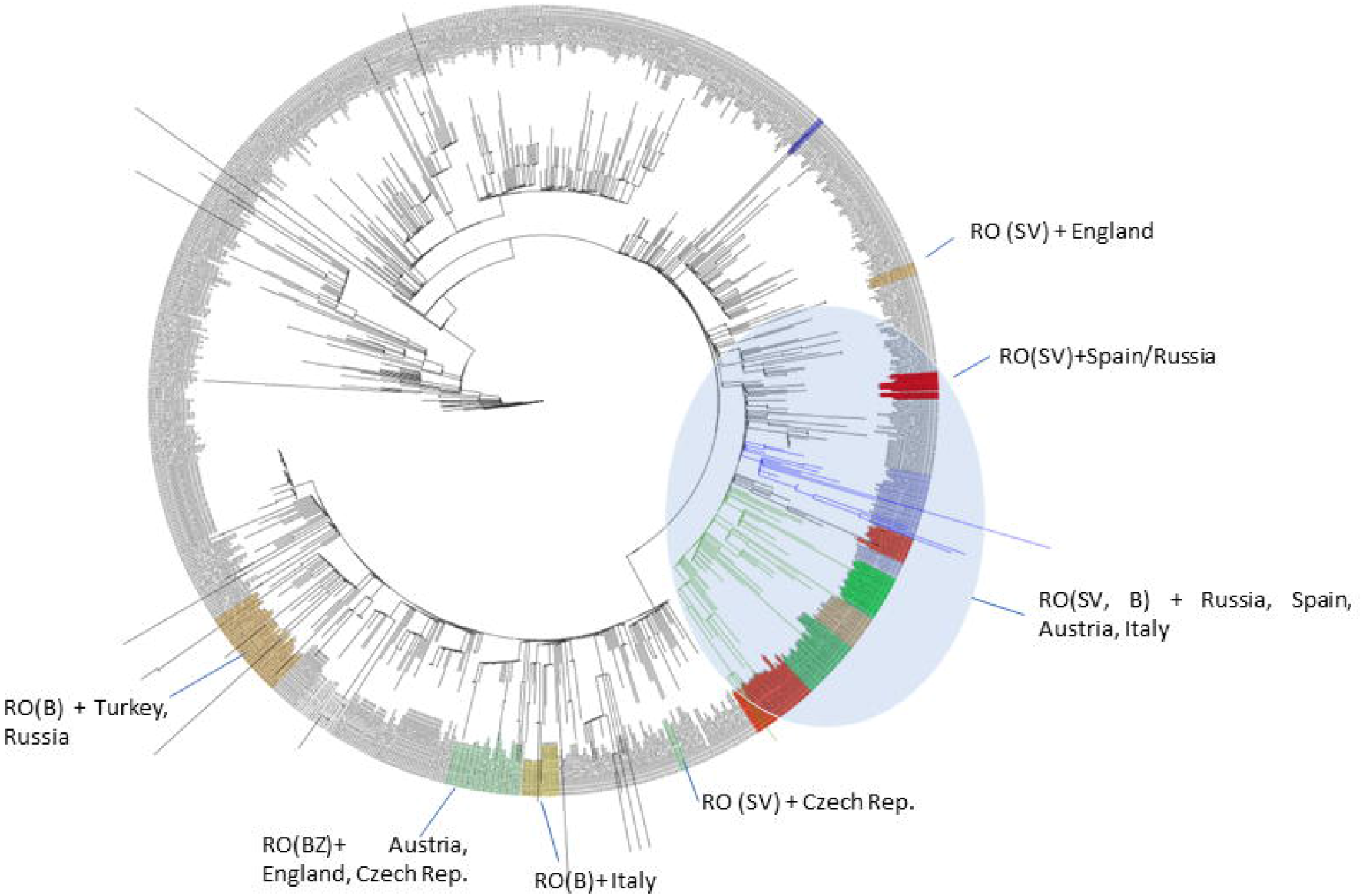
Phylogenetic distribution of SARS-CoV-2 emergence in Romania in relation to 859 European accessions. Importantly, the Romanian phylogenetic clustering was mostly preserved in the larger European phylogenetic analysis. Approximately one third of Romanian strains, including over 30 samples from Suceava region and 4 samples from Bucharest, clustered together with samples from Russia, Spain and Austria. This cluster had two distinct subclusters that corresponded with particular sample collection dates. One subcluster comprised earlier samples (March and April 2020), grouped together with strains from Russia, Turkey and Italy while the second subcluster that comprised Romanian samples from May and June 2020, were mainly grouped with samples from France, Spain, Russia, Scotland and Belgium. A second cluster consisted of 6 Bucharest strains sampled between 12 and 18 April, 2020 that tracked together exclusively with a set of strains from Italy (24 March - 9 April, 2020). A third cluster, also comprising strains from the southern part of the country (Buzau) collected in June, grouped with strains from Austria, England and Czech Republic. Another strain from Suceava (April, 2020) was related to samples from the Czech Republic (March), while a sample from Buzau (June) was related to samples from England (May).

The relationship between strains was also obtained when inferring phylogenies using Nextstrain application, which is based on more than 30.000 sequences from the GISAID database (Fig. 8).

**Fig 8.**
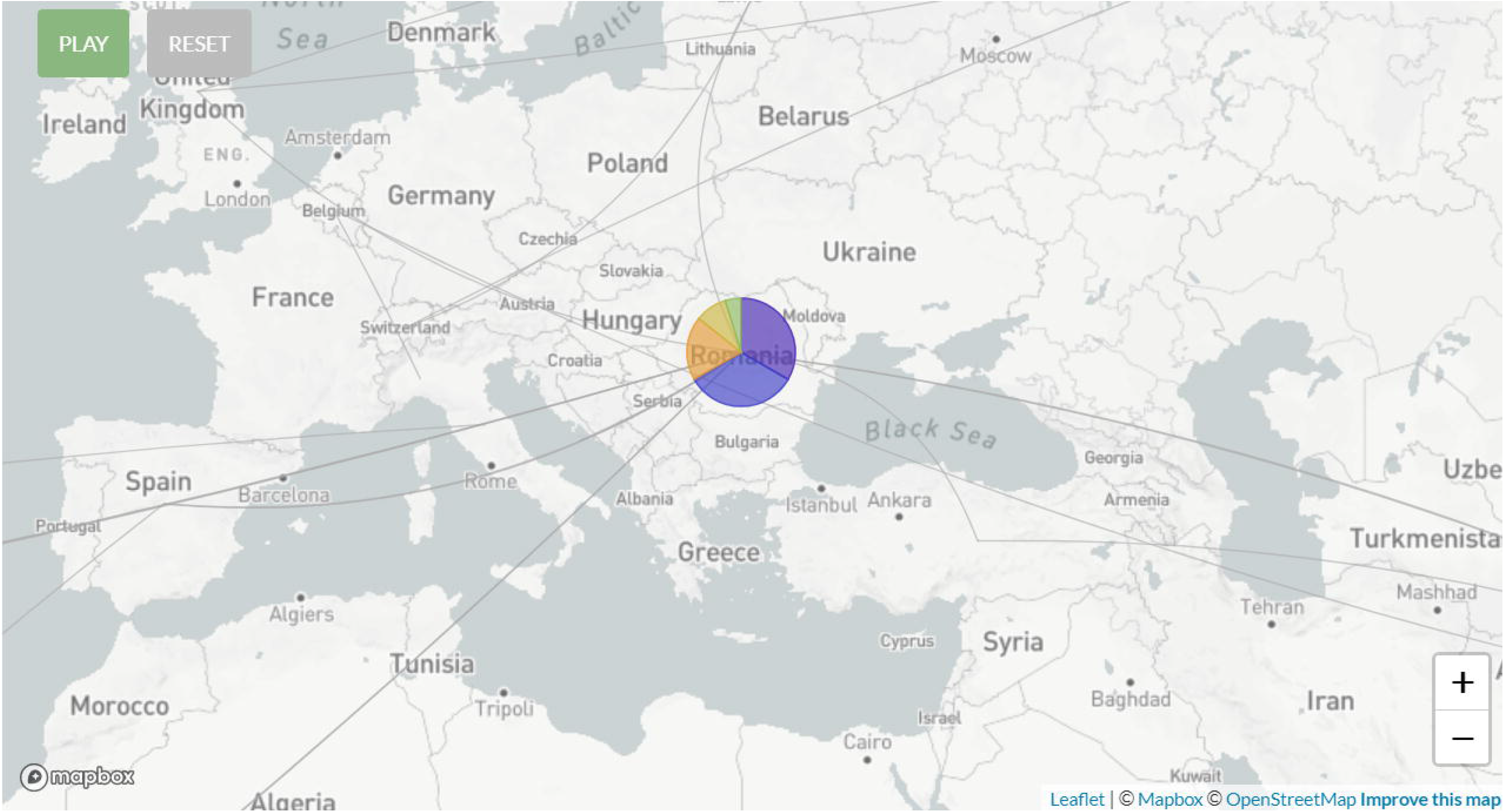
Presumptive routes of transmission of SARS-CoV-2 from Suceava (generated from nextstrain.org, based on available European strains, route lines corresponding to simulation in August 2020). As noted, traces of strain movement across Romania originated mainly from Spain, Italy, Greece, Turkey, UK and far East of Russia. With respect to GISAID pangolin lineages, most of our strains from Suceava (n=57) belong to the B.1.5 lineage, GISAID G clade, while 4 strains belong to the B.1.1 lineage, GR clade and 1 strain to the B.1, G clade. When taking into account the European samples subset provided by Nextstrain, 28 of our samples from Suceava belong to 20A Nextstrain clade, while 2 belong to the 20B clade (Fig. 9). The temporal dynamic within analysed samples was confirmed through a Bayesian inferrence of strains, in order to calculate mean time to most recent ancestor (tMRCA). The date obtained was November 2019, using both a HKY and a GTR + gamma substitution model, confirming the temporal signal (0.954 correlation coefficient, 639 Estimated Sample Size – ESS) present in our analyzed strains. A total 112 Romanian samples in coloured areas, indicated with RO; SV, Suceava; B, Bucharest; BZ, Buzau. Large blue oval cluster: 36 Romanian samples, with subclusters; listed countries indicate closest origin with Romanian strains.

**Fig 9.**
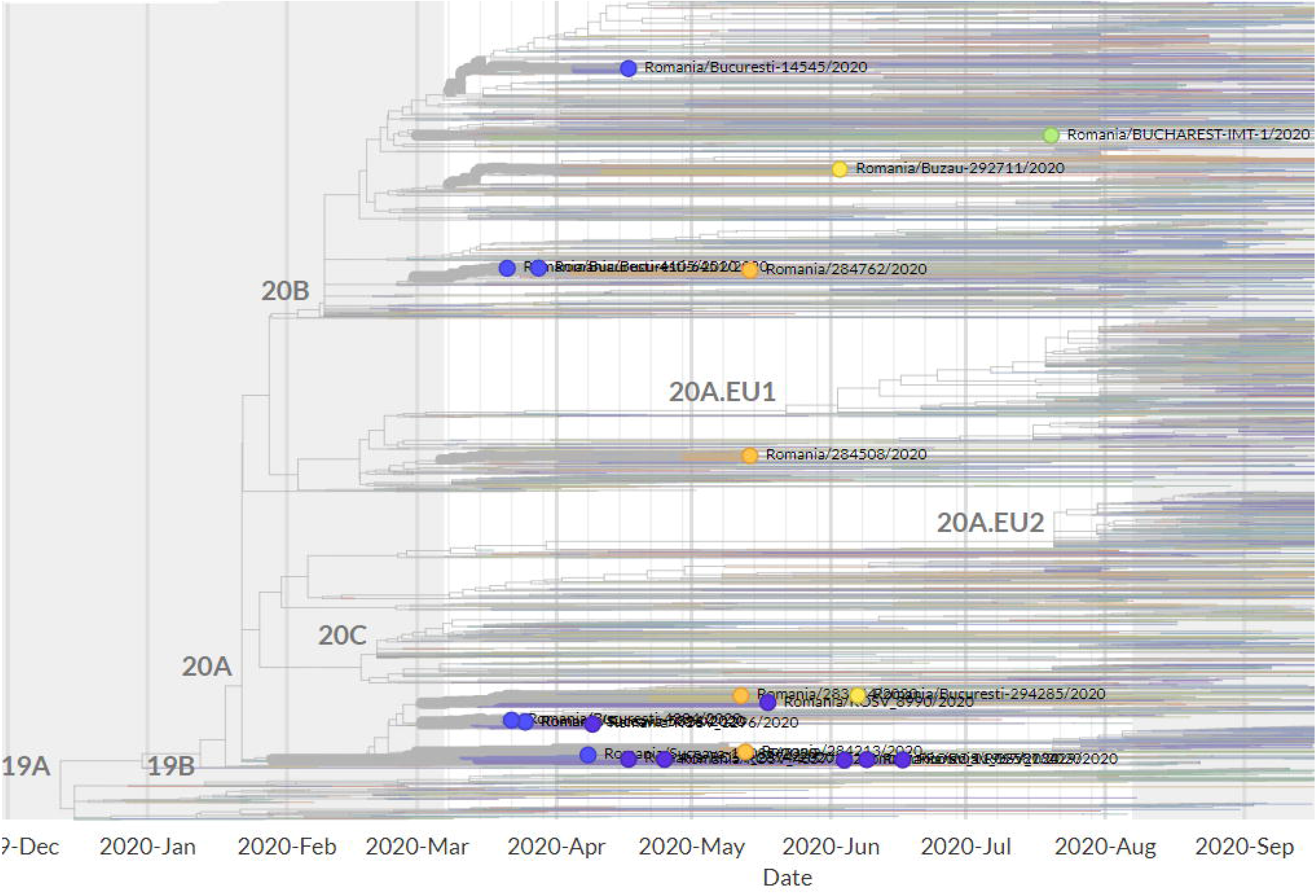
Phylogenetic analysis of SARS-CoV-2 emergence in Suceava and other Romanian regions within Nextstrain phylogeny/clades. Large purple dots, Suceava, on the left of the tree and other Romanian regions within Nextstrain phylogeny/clades.

### Clinical parameters and outcomes

Of the 62 patients from whom viral sequences were obtained, 23 were females, and 39 were males (average 54 years). With respect to the severity of the disease, 6 patients were asymptomatic, 35 had a mild condition, and 21 displayed a severe form. The hospitalisation length ranged between 4 and 39 days (mean 20 days). In order to identify factors that significantly influenced disease severity and clinical outcome we examined viral copy number, number of mutations, number of hospitalized days, age, sex and certain comorbidities, such as diabetes, obesity and hypertension. Among continuous variables, only the number of hospitalized days showed significant differences between asymptomatic and mild status patients (Table 3). Although the average number of hospitalized days was highest in mildly diseased patients, the hospitalization days ranged from 4 to 22 days for asymptomatics, 7 to 39 days for mild status and 9 to 34 days for severe status. Sex was not significantly correlated with the severity of disease (Pearson’s rho = 0.17).

**Table 3.** Sample and patient related factors

| Factor | Asymptomatic | Mild | Severe |
| --- | --- | --- | --- |
| *Viral copies* | 6.05E+04<sup>a</sup>±5.23E+04 | 8.86E+04<sup>a</sup>±2.50E+04 | 8.40E+04<sup>a</sup>±3.49E+04 |
| *Hospitalized days* | 12.40<sup>a</sup>±3.43 | 23.39<sup>b</sup>±1.76 | 18.41<sup>ab</sup>±1.91 |
| *Age* | 57.83<sup>a</sup>±3.98 | 49.80<sup>a</sup>±3.47 | 55.10<sup>a</sup>±3.79 |
| *Number of mutations* | 8.00<sup>a</sup>±0.58 | 9.74<sup>a</sup>±0.81 | 9.33<sup>a</sup>±0.61 |

Superscripts with different letters on the same row indicate statistical significance ( $p < 0.05$ ).

When examining the effect of comorbidities on clinical outcome, the percentages of hypertensive, diabetic and obese patients were higher among mild and severe forms (Table 4). When comparing asymptomatic patients with mild and severe forms, logistic regression analysis revealed a significant association with hypertension and diabetes with an odds ratio (risk ratio) of 8.47 (p=0.008) and 3.28 (p=0.037) for hypertension and diabetes, respectively. These patients were eight (hypertension) and three (diabetes) times more likely to develop a severe form of the disease.

**Table 4.** Percentage (%) of patients with comorbidities

| Disease severity | Hypertension | Obesity | Diabetes |
| --- | --- | --- | --- |
| Asymptomatic | - | 16.67 | - |
| Mild | 37.14 | 34.29 | 17.14 |
| Severe | 42.86 | 33.33 | 23.81 |

None of the above factors (viral copy number, days of hospitalisation, comorbidities) were significantly associated with the number of deaths possibly due to the low numbers of such patients (n=7). It should be noted however, that out of the 7 deceased patients, 2 had hypertension, 1 was diabetic and 3 were obese.

To investigate possible effects of viral mutations on disease outcome we screened for non-synonymous reccuring mutations, in patients with severe clinical forms and those deceased. We found that 40% of the severe cases had a mutation corresponding to position 27707 of the ORF7a region and three patients presented with a mutation at position 3225 which is assigned to ORF1ab region. In the deceased patients, recurring mutations were identified at position 27707 (3 samples), 3225 (3 samples) and 29725, corresponding to the spike gene of the virus (2 samples) (Fig. 10)

**Fig 10.**
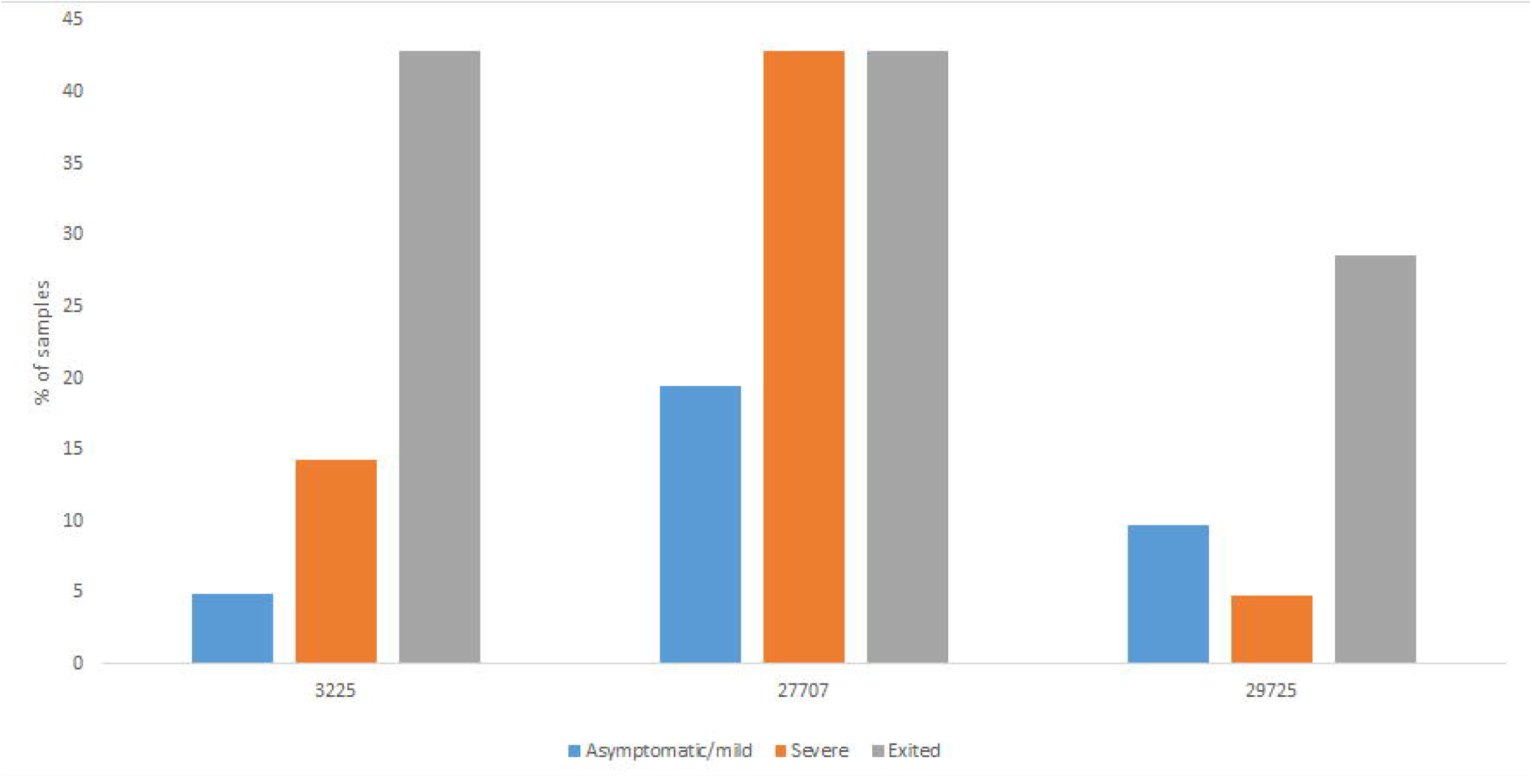
Percentages of frequent recurrent mutations encountered in different patient groups (n>3).

### Viral protein modelling

To assess the putative effect of mutations on viral structure, proteins corresponding to mutated regions were modelled accordingly. Mutation at position 27707 (C>T) altered the ORF7a protein, 121 aminoacid residues long, changing the alanine in position 105 to a valine. Considering that analytical structural data is only available for the 16-82 residues range, protein modelling was performed *ab initio*, independently, using Phyre2 (17) and I-TASSER (18), for both mutated and wild proteins (Fig. 11). Superimposing structures yielded a RMSD value of 2.33 Å for Phyre2 predicted structures and 2.29 Å for I-TASSER ones and, correspondingly, a TM score of 0.85 for both cases.

**Fig 11.**
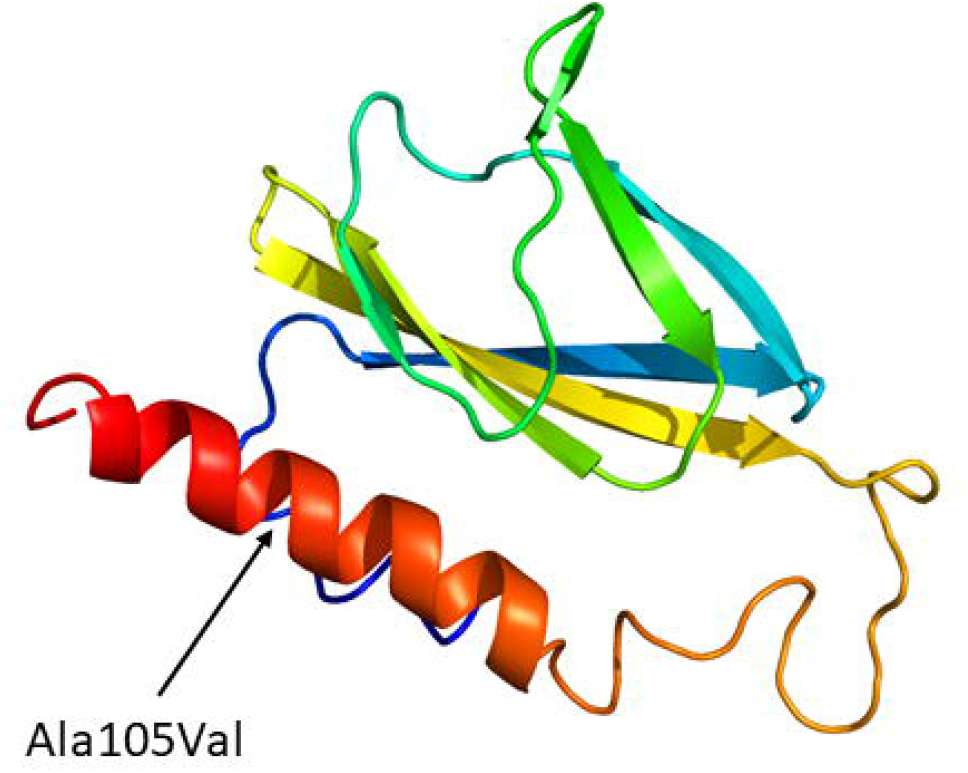
abinitio model of entire ORF7a protein.

**Fig 12.**
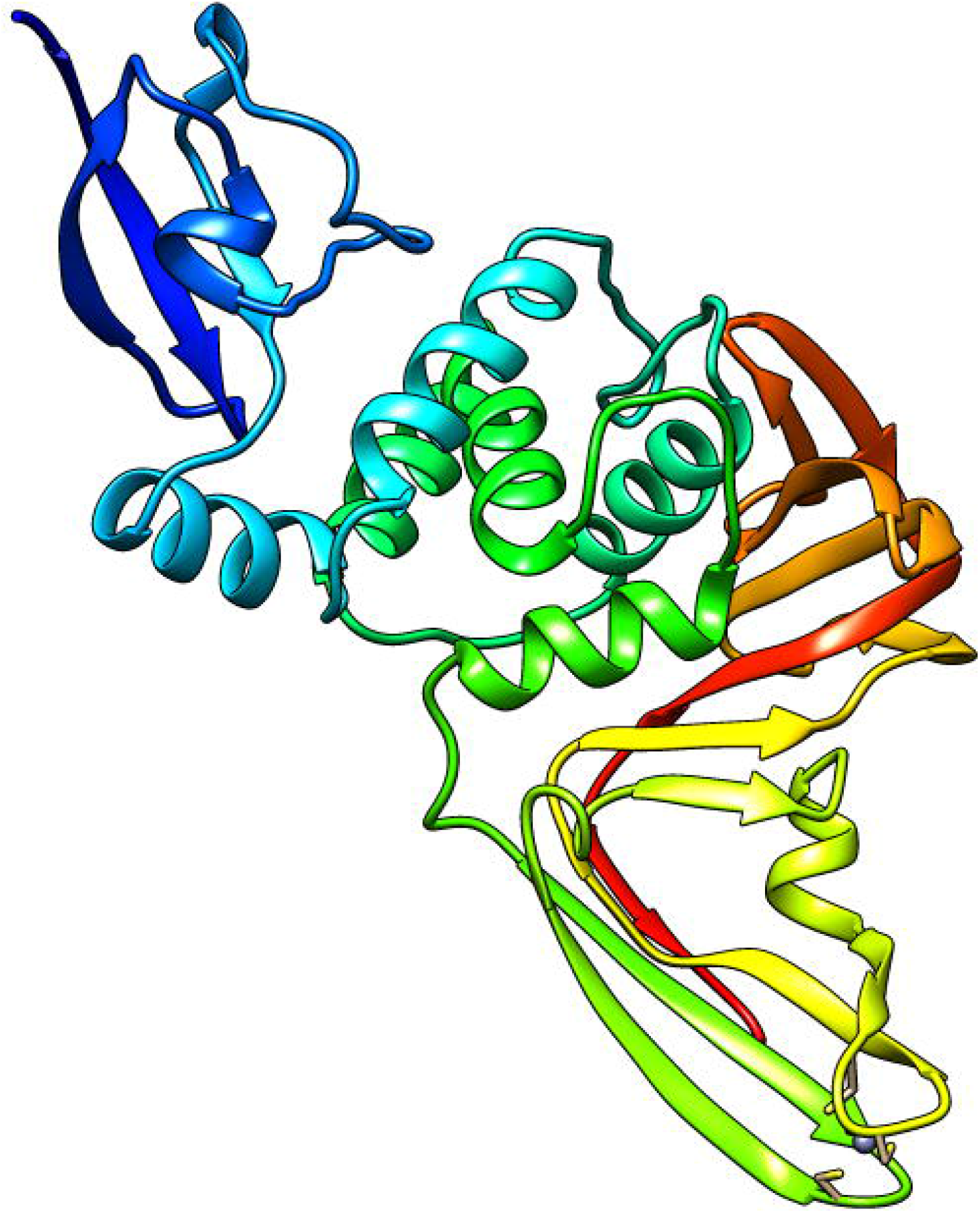
abinitio model of NSPNSP3a.

Another mutation was represented by a change from cytosine to adenine at position 3225 C>A, leading to a substitution of threonine by asparagine at position 987 of the ORF1ab region. This mutation appeared in the papain-like protease (NSPNSP3) coding region, at position 169, domain NSPNSP3a. This region is described mainly for SARS-CoV-1 and consists of two subdomains, an ubiquitin-like domain (Ubl1) and a Glu-rich, acidic region, located at the N-terminal of NSP3. For SARS-CoV-2, no structures are available for this region, thus after alignment with the similar domain of SARS-CoV-1, the first 205 residues of NSPNSP3 were *ab initio* modeled. The two models, for mutated and original sequence, recorded a RMSD of 3.81 Å and a TM-score of 0.60 (Fig. 12).

## Discussion

The sequence and phylogenetic analyses of SARS-CoV-2 strains showed that Romanian samples appear heterogenous in the quality and quantity of mutations and supports the idea of multiple sources of virus introduction in the country. When integrating sequencing data with clinical parameters of patients, our results identified specific viral mutations that were associated with the degree of disease severity and patients’ comorbidities.

### Characteristics and introduction of SARS-CoV-2 strains in Romania

Among detected single nucleotide polymorphisms, there was a high, 3:1 ratio of transitions to transversions. This ratio was described in SARS-CoV-2 mutations, where, supossedly, cytosine deaminases may be responsible for C>T changes, while G>T ones may be the result of oxo-guanine arising from reactive oxygen species [19]. The SNP profile of our sequenced samples, is in line with the large picture observed in over 40,000 worldwide samples regarding the prevalence of C>T mutations [20]. However, secondly abundant in our samples, was the G>T transversion, as compared to A>G transition, which ranks third. This higher C>T numbers might be explained by the differential activity of viral RNA editing enzymes. For example, APOBECs (Apolipoprotein B mRNA editing complex) and ADARs (Adenosine Deaminases Acting on RNA), generate such mutations on positive strand RNA, pointing to the abundance of this form or viral RNA, over negative strand RNA within samples [21].

All 62 samples analyzed from Suceava share a set of mutation common to European samples. Specifically, mutations at positions 241 (c.-25C>T), 3037 (c.2772C>T), 14408 (c.14143C>T), 20268 (c.20003A>G) and 23403 (c.1841A>G) are frequent in European samples [12] and were identified early in the pandemic evolution, as a signature for one of the superspreaders that originated from Wuhan (Yang et al., 2020). Among the most abundant observed mutations, 241C > T belongs to the leader sequence, with significance for discontinuous sub-genomic replication. This mutation co-evolved with other three major mutations, 3037C > T, 14408C > T, and 23403A > G. As a consequence, a synonymous mutations in NSP3, a P323L mutation in RNA primase, and a D614G mutation in spike glycoprotein occurred. The combination of these four mutations was mostly observed in European strains, and is associated with more severe forms of infection and, possibly, with increased transmissibility [22]. Interestingly, the signature of another superspreader, which was less encountered in European strains, namely the change in G11083T was present in two of our samples. This mutation was observed in samples from France and China [23].

Mutation A20268G was recorded in samples collected before 26 of May, in 45 % of strains from Spain [24], and was also present in most of our samples from Suceava. However, 7 of our 62 sequenced samples did not share this mutation, including those belonging to B.1 and B.1.1 lineages but not the B.1.5. The strain belonging to B.1 lineage, sampled on June 5, was close in our phylogenetic analysis, with another Romanian sample from Iasi and, overall, with samples from France and Belgium, in a distinct subcluster. Two other B.1.1 lineage strains, from June 8 and 9, were grouped, along with samples from Russia (Moscow, St. Petersburg, Novosibirsk), England, Sweden, Austria, Croatia, Bosnia and Herzegovina. A third B.1.1 lineage strain, from June 8 was, again, in a distinct sublcuster, with samples from Turkey and Greece, while the fourth B.1.1 strain, from April 16, was closer to Czech Republic strains. When examining sampling dates for the 7 strains without A20268G mutation, it appears that they were introduced by a patient in Suceava around April 16, possibly from the Czech Republic. The other 5 strains sampled between June 5 and 19 could be the result of local transmission or introduction from other countries specified above. All other samples from Suceava were grouped in the same large cluster, along with strains from several European countries.

When examining Romanian samples belonging to B.1.1 lineage from outside Suceava, we found 6 strains from Bucharest sampled between 12 and 18 of April, that were distinctly grouped with Italian samples dated March-April. Another group of 11 strains from Bucharest and Mioveni, sampled between 21 March and 14 May were grouped with strains from Turkey and Russia. Three other southern Romanian strains from Buzau, dated early June, grouped with March and April samples from Austria, Czech Republic, England and Poland. Therefore, It appears that B1.1. strains originated in Romania around late March to early April from England, Austria and Czech Republic. Otherwise, Romanian samples belonging to B.1.5 lineage were mixed, mainly with samples from Russia, Italy, Spain and Austria. These results support a multiple source of virus introduction in the country with one route being Spain, Italy and Turkey, the second England, Austria and Czech Republic and a third from East Russia and beyond. Clearly, strains from Suceava grouped together, perhaps because the city and adjacent regions were under mobility restrictions, while a wider distribution of the virus was observed across Romania, especially after restrictions were removed.

### COVID-19 disease and patient status

Examination of COVID-19 patient data from Suceava showed a high variability in clinical manifestation and disease severity. Our study showed a significant increase in disease severity in patients with hypertention and diabetes. This is in line with previous work as well as with results from a recent study in Romanian COVID-19 patients indicating risk ratios (RR) with values of 6.4 for diabetes and 3.3 for hypertension [25–27]. Currently, there is not a consensus on the mechanisms by which hypertension or diabetes increase morbidity risk in COVID-19 patients. ACE-1 inhibitors and angiotensin II receptor blockers, that are used for diabetics and hypertensives, upregulate ACE-2 expression, the SARS-CoV-2 receptor. While this may have protective effects against lung injury, it increases the chances of acquiring the disease. Likewise, a hyperglycemic environment increases virulence of some pathogens, phagocytosis, chemotaxis, response of T cells and neutrophils, decreased immune response while production of interleukins is restricted [28]. Furthermore, coronaviruses may increase glycemic levels by damaging pancreatic islet cells [29]. ACE-2 expression polymorphism present in humans could influence both the susceptibility and outcome of COVID-19 [30]. Finally, in our patients, age was not related with disease severity, outcome or length of hospitalization, altough age was considered a risk factor in Italy [31] and might be a driver for disease trajectory [32].

### Viral protein modifications

Among mutations in SARS-CoV-2, some result in protein structure and properties changes. One such mutation is D614G, which, over the course of one month, became prevalent in viral strains worldwide. Although D614G mutation was associated with lower Ct values in RT-PCR analyses [11], our samples recorded a very large range of Cts, between 17 and 36, suggesting that several factors, probably both virus and host related, influence viral titres.This mutation is associated with less shedding of S1 subdomain of the S protein, increased viral stability and transmission, although not necessarily with increased disease severity [33], while other studies associated D614G and P4715L with increased mortality.

Another nonsynonymous recurrent mutation in a large number of our samples was the replacement of alanine with valine, in position 105. Our model of protein modification showed that this mutation has a high probability to modify an alfa-helix stretch into a beta-sheet conformer since valine is a hydrophobic amino acid known to be one of the best β-sheet former [34]. Therefore, the replacement of alanine 105 residue to valine favors β-sheet’s secondary structure. The ORF7a region in genomes of coronaviruses encodes a 5.5 kDa protein, with a putative role in enhancing virulence in SARS-CoV. In SARS-CoV-1, the 122 residues long protein (accessory protein 7a) an integral membrane protein is, localized in the Golgi compartments, probably in the budding regions [35]. It was shown to be involved in cell apoptosis, through caspase-dependent pathway, cell protein synthesis inhibition, cell cycle progression blockage and proinflammatory action, thus altering the host cellular environment [36, 37]. Interestingly, this protein is known to interact with a viral release inhibitor, bone marrow stromal antigen 2 (BST-2 or CD317 or tetherin). BST-2 is an interferon-inducible factor that tethers various nascent enveloped viruses on the cell surface, playing an important role in viral infection [38]. The antiviral function of BST-2, in the case of SARS-CoV, occurs by stopping the egression of virions through the plasma membrane and inhibits glycosylation of BST-2 while removes its antiviral function [39]. Although ORF7a was proposed as a potential candidate for antiviral drug development [40], other groups have reported significant mutations in ORF7a [41].

Another recurrent mutations in our samples, associated with a large proportion of severe disease instances, was the threonine to asparagine replacement in NSP3a gene. The mutation that occur in the structurally disordered domain, involved the amino acid residue 169 and constituted in a Thr →Asn amino acid substitution. NSP3a, is one of the seven domains of SARS-CoV NSP3 polypeptide known to be involved in RNA replication [42] and consists of a 112-residue N-terminal subdomain with a homogeneous content of amino acids and a C-terminal subdomain rich in acidic residues. NMR studies revealed that the subdomain NSP3a(1-112) exhibits a globular ubiquitin-like fold with two additional helices while the Glu-rich acidic domain (residues 113 to 183), also called “hypervariable region”, was shown to be structurally disordered [43]. These unique structural elements are involved in interactions with single-stranded RNA. Structural similarities with proteins involved in various cell-signaling pathways indicate possible roles of NSP3a in viral infection and persistence [43]. The function of the glutamic acid-rich region is still not known, however similar Glu-rich region were observed in the transcription factor Mytl1l known to be involved in the general function of binding nucleic acids. NSP3a can be classified as a “low complexity region”, found in many viruses, including Coronaviridae. Such regions are considered to be highly immunogenic and, importantly, they share high simmilarity with human epitopes, thus making them a risk for antiviral drug or testing development [44]. The enrichment of glutamic acid was found as a feature of the highly immunogenic polypeptides, in other organisms as well [45].

## Conclusions

COVID-19 became an ubiquitary presence in Romania, affecting all groups of individuals regardless of age, sex or other factors. The SARS-CoV-2 sources that triggerred the first largest outbreak of COVID-19 in Romania could be mainly traced to Spain, Italy and Russia, with fewer strains related to those from Czech Republic, Belgium and France. Specific mutations in the genome regions ORF1a, S and ORF7 were identified that were associated with the severity of the disease and comorbidities such as hypertention and diabetes. Further viral genomic analyses evolution is critical for detection of mutations, virus containment and timely treatment.

## Methods

### Sample collection

Viral RNA was obtained from samples of patients hospitalized in the Suceava County Regional Hospital, collected between 10.04.2020 and 19.06.2020. Patients signed informed consent for data access and the study was approved by the University of Suceava Research Ethics Committee. Criteria for patient selection included age, sex, severity of the disease, number of days in hospital and existing comorbidities. Samples were collected by nasopharingeal swabs from patients presenting with COVID-19-like symptoms. Clinical, epidemiological and demografic data were taken from patients’ medical records.

### Sample preparation and sequencing

RNA extraction was performed using Bioneer AccuPrep® Viral RNA Extraction Kit. RNA extracts were evaluated for viral copy numbers (TaqMan 2019-nCoV Assay Kit v1, Applied Biosystems, USA), and a number of SARS-CoV-2 positive samples were selected for analysis. RNA (100 ng) was reverse transcribed using SuperScript™ VILO™ cDNA Synthesis Kit (Invitrogen, USA), according to product protocol. Targets for sequencing were obtained based on Ion AmpliSeq™ SARS-CoV-2 Panel (ThermoFisher, USA). Library preparation was made using Ion AmpliSeq™ Library Kit Plus (ThermoFisher, USA), then libraries were loaded on sequencing chips using Ion Chef equipment. Next generation sequencing was performed on Ion S5 Gene Studio, using Ion Torrent 540 chips.

### Sequence processing and data availability

Sequences were assembled using the Iterative Refinement Meta Assembler (IRMA), after which variants were called with Torrent VariantCaller plugin, referenced to the Wuhan SARS-CoV-2 sequence and annotated using SnpEff plugin. Sequences were uploaded in GISAID database on 2020-07-11 and 2020-07-16.

### Phylogenetic analysis

To assess phylogenetic placement of Romanian strains that included those from Suceava and other regions, a GISAID survey was performed, selecting European, and UK strains, under the “high coverage”, “low coverage excluded”, “complete” criteria. All 112 Romanian samples present in GISAID at the time of writing this article were included, together with another 1016 European samples. After removing duplicate sequences and those with long stretches of “*nnn*”, a total of 859 strains were considered for analyses. Prior to phylogenetic analyses, all samples were aligned using MAFFT algorithm, then trimmed at ends, to remove unnecessary artifacts caused by sequencing in those areas.

Phylogenetic analysis of our sequenced samples and selected European ones was performed based on maximum likelihood algorithms, using RaxML-HPC2 workflow on CIPRES platform, phylo.org [46]. Set parameters were GTR Gamma model and a bootstrap value of 100. The resulting tree was loaded, visualized and annotated in ITOL platform, itol.embl.de [47]. Bayesian time dated phylogenetic analysis of the data set was performed using BEAST 2.6.3, with Beagle library enabled. A HKY + Γ model of nucleotide substitution and a strict clock were assigned, using a coalescent exponential population model. A continuous-time Markov chain was employed, posterior distributions of parameters were estimated by sampling every 5000 steps over a total of 50 million MCMC steps. Each analysis was run in duplicate to check for convergence, and the first 10% of samples were discarded as burn-in. Sampling was considered effective when a minimum of 200 Estimated Sample Size was reached.

## Data Availability

Data submitted to GISAID https://www.gisaid.org/ For Accesion info, refer to S1 Table

## Statistical analyses

Differences between patient groups (asymptomatic, mild, severe) were assessed using ANOVA, followed by a Duncan post-hoc test, while correlations were evaluated by Spearman’s coefficient. Ordinal or binary logistic regression was applied for severity categories, while binary logistic regression was applied when various factors were assessed in relation with patient’s status. All statistical analyses were conducted using Minitab 19.2020.1 (Minitab LLC).

## List of abbreviations

Asn: Asparagine
GISAID: GISAID EpiFlu™ Database and Data, https://www.gisaid.org/
IL-8: Interleukin 8
kb: kilobases
MAP: mitogen activated protein kinase
NF-êB: Nuclear factor – kappa B
NSP3a: Non-structural protein 3, subdomain 3a
ORF: Open reading frame
Thr: Threonine

## Supporting information

S1 Table. Accession number of Romanian/Suceava SARS-CoV-2 genome sequences submitted to GISAID

