## Supplemental Table 1 for "Analysis of genome characteristics and transmission of SARS-CoV-2 strains in North-East of Romania during the first COVID-19 outbreak"

**S1 Table. Accession number of Romanian/Suceava SARS-CoV-2 genome sequences submitted to GISAID**

| No. | Virus name | Accession ID | Collection date |
|---|---|---|---|
| 1 | hCoV-19/Romania/Suceava-4682/2020 | EPI_ISL_468137 | 22/03/2020 |
| 2 | hCoV-19/Romania/Suceava-5709/2020 | EPI_ISL_468138 | 25/03/2020 |
| 3 | hCoV-19/Romania/Suceava-5710/2020 | EPI_ISL_468139 | 25/03/2020 |
| 4 | hCoV-19/Romania/Suceava-5722/2020 | EPI_ISL_468140 | 25/03/2020 |
| 5 | hCoV-19/Romania/Suceava-5806/2020 | EPI_ISL_468141 | 25/03/2020 |
| 6 | hCoV-19/Romania/Suceava-5816/2020 | EPI_ISL_468142 | 25/03/2020 |
| 7 | hCoV-19/Romania/Suceava-5893/2020 | EPI_ISL_468143 | 25/03/2020 |
| 8 | hCoV-19/Romania/Suceava-5894/2020 | EPI_ISL_468144 | 25/03/2020 |
| 9 | hCoV-19/Romania/Suceava-11085/2020 | EPI_ISL_468146 | 08/04/2020 |
| 10 | hCoV-19/Romania/Suceava-31645/2020 | EPI_ISL_468157 | 29/05/2020 |
| 11 | hCoV-19/Romania/Suceava-31646/2020 | EPI_ISL_468158 | 29/05/2020 |
| 12 | hCoV-19/Romania/Iasi-296512/2020 | EPI_ISL_471421 | 10/06/2020 |
| 13 | hCoV-19/Romania/ROSV-2/2020 | EPI_ISL_486834 | 25/05/2020 |
| 14 | hCoV-19/Romania/ROSV_12646/2020 | EPI_ISL_486854 | 10/06/2020 |
| 15 | hCoV-19/Romania/ROSV/2020 | EPI_ISL_486855 | 08/06/2020 |
| 16 | hCoV-19/Romania/ROSV_12287/2020 | EPI_ISL_486856 | 2020 |
| 17 | hCoV-19/Romania/ROSV_10626/2020 | EPI_ISL_491036 | 26/05/2020 |
| 18 | hCoV-19/Romania/ROSV_11161/2020 | EPI_ISL_491037 | 29/05/2020 |
| 19 | hCoV-19/Romania/ROSV_11967/2020 | EPI_ISL_491038 | 04/06/2020 |
| 20 | hCoV-19/Romania/ROSV_11990/2020 | EPI_ISL_491039 | 05/06/2020 |
| 21 | hCoV-19/Romania/ROSV_12235/2020 | EPI_ISL_491040 | 08/06/2020 |
| 22 | hCoV-19/Romania/ROSV_12278/2020 | EPI_ISL_491041 | 07/06/2020 |
| 23 | hCoV-19/Romania/ROSV_12421/2020 | EPI_ISL_491042 | 08/06/2020 |
| 24 | hCoV-19/Romania/ROSV_12442/2020 | EPI_ISL_491043 | 08/06/2020 |
| 25 | hCoV-19/Romania/ROSV_12587/2020 | EPI_ISL_491044 | 09/06/2020 |
| 26 | hCoV-19/Romania/ROSV_12615/2020 | EPI_ISL_491045 | 09/06/2020 |
| 27 | hCoV-19/Romania/ROSV_12618/2020 | EPI_ISL_491046 | 09/06/2020 |
| 28 | hCoV-19/Romania/ROSV_12619/2020 | EPI_ISL_491047 | 09/06/2020 |
| 29 | hCoV-19/Romania/ROSV_12723/2020 | EPI_ISL_491048 | 10/06/2020 |
| 30 | hCoV-19/Romania/ROSV_12812/2020 | EPI_ISL_491049 | 11/06/2020 |
| 31 | hCoV-19/Romania/ROSV_1296/2020 | EPI_ISL_491050 | 09/04/2020 |
| 32 | hCoV-19/Romania/ROSV_13109/2020 | EPI_ISL_491051 | 15/06/2020 |
| 33 | hCoV-19/Romania/ROSV_1332/2020 | EPI_ISL_491052 | 09/04/2020 |
| 34 | hCoV-19/Romania/ROSV_13429/2020 | EPI_ISL_491053 | 17/06/2020 |
| 35 | hCoV-19/Romania/ROSV_1345/2020 | EPI_ISL_491054 | 09/04/2020 |
| 36 | hCoV-19/Romania/ROSV_1837/2020 | EPI_ISL_491055 | 11/04/2020 |
| 37 | hCoV-19/Romania/ROSV_1979/2020 | EPI_ISL_491056 | 11/04/2020 |
| 38 | hCoV-19/Romania/ROSV_2113/2020 | EPI_ISL_491057 | 11/04/2020 |
| 39 | hCoV-19/Romania/ROSV_2492/2020 | EPI_ISL_491058 | 16/04/2020 |
| 40 | hCoV-19/Romania/ROSV_2778/2020 | EPI_ISL_491059 | 16/04/2020 |
| 41 | hCoV-19/Romania/ROSV_2794/2020 | EPI_ISL_491060 | 16/04/2020 |
| 42 | hCoV-19/Romania/ROSV_2873/2020 | EPI_ISL_491061 | 17/04/2020 |
| 43 | hCoV-19/Romania/ROSV_2874/2020 | EPI_ISL_491062 | 17/04/2020 |
| 44 | hCoV-19/Romania/ROSV_2899/2020 | EPI_ISL_491063 | 17/04/2020 |
| 45 | hCoV-19/Romania/ROSV_3019/2020 | EPI_ISL_491064 | 17/04/2020 |
| 46 | hCoV-19/Romania/ROSV_3169/2020 | EPI_ISL_491065 | 18/04/2020 |

| | | | |
|---|---|---|---|
| 47 | hCoV-19/Romania/ROSV_3194/2020 | EPI_ISL_491066 | 19/04/2020 |
| 48 | hCoV-19/Romania/ROSV_3242/2020 | EPI_ISL_491067 | 19/04/2020 |
| 49 | hCoV-19/Romania/ROSV_3316/2020 | EPI_ISL_491068 | 20/04/2020 |
| 50 | hCoV-19/Romania/ROSV_3376/2020 | EPI_ISL_491069 | 20/04/2020 |
| 51 | hCoV-19/Romania/ROSV_3561/2020 | EPI_ISL_491070 | 21/04/2020 |
| 52 | hCoV-19/Romania/ROSV_3960/2020 | EPI_ISL_491071 | 22/04/2020 |
| 53 | hCoV-19/Romania/ROSV_3962/2020 | EPI_ISL_491072 | 22/04/2020 |
| 54 | hCoV-19/Romania/ROSV_4271/2020 | EPI_ISL_491073 | 23/04/2020 |
| 55 | hCoV-19/Romania/ROSV_4342/2020 | EPI_ISL_491074 | 24/04/2020 |
| 56 | hCoV-19/Romania/ROSV_4657/2020 | EPI_ISL_491075 | 25/04/2020 |
| 57 | hCoV-19/Romania/ROSV_4659/2020 | EPI_ISL_491076 | 25/04/2020 |
| 58 | hCoV-19/Romania/ROSV_4666/2020 | EPI_ISL_491077 | 25/04/2020 |
| 59 | hCoV-19/Romania/ROSV_4714/2020 | EPI_ISL_491078 | 26/04/2020 |
| 60 | hCoV-19/Romania/ROSV_4730/2020 | EPI_ISL_491079 | 26/04/2020 |
| 61 | hCoV-19/Romania/ROSV-1815/2020 | EPI_ISL_678398 | 11/04/2020 |
| 62 | hCoV-19/Romania/ROSV-10281/2020 | EPI_ISL_678397 | 25/04/2020 |
